# “*You become free, you can tell her anything*”: Perceptions of a peer-based medication delivery implementation strategy to improve hypertension medication adherence in western Kenya

**DOI:** 10.64898/2026.03.05.26347760

**Authors:** Caroline Watiri, Juddy Wachira, Benson Njuguna, Jessica Gjonaj, Kibet Kangogo, Mercy Korir, Jeremiah Laktabai, Imran Manji, Sonak D Pastakia, Dan N. Tran, Rajesh Vedanthan

## Abstract

**Background:** In low- and middle-income countries, the burden of hypertension is increasing. Medication adherence is a critical component of reducing hypertension-related cardiovascular disease (CVD) risk and death. There are many barriers to hypertension medication adherence, including challenges with access to and possession of medication. To address these challenges, we aim to implement a strategy in rural western Kenya that combines peer delivery of medications and health information technology to improve hypertension medication possession and adherence. Recognizing that stakeholder experience and knowledge can be useful to optimize successful implementation, we sought to assess micro- and macro-level stakeholder perceptions of the planned implementation strategy.

**Methods:** Focus group discussions in both English and Kiswahili were conducted among people living with hypertension, community members, and health workers. In addition, key informant interviews were conducted with public sector health administrators including the program/policy planners for non-communicable diseases at the national and county levels. Content analysis of all transcripts was conducted. A codebook containing deductive codes was generated based on *a priori* themes identified from the interview guide. These included the perceptions of peers being involved in health service provision, medication delivery, psychosocial support, and the use of health information technology. Emerging themes were also identified and integrated into the results. The investigator team pooled codes according to conceptual alignment and integrated them into common themes after joint review and discussion. NVIVO 12 was used for the data analysis.

**Results:** The PT4A implementation strategy was perceived to have both benefits and potential challenges. Major themes included the importance of trust resulting from a safe space to share experiences with peers, increased access to medications, improved hypertension management at the facility and community levels, and anticipated improved health outcomes for people living with hypertension. The success of the program was felt to rely heavily on the peers’ competency and how well they communicated, which was viewed as a potential challenge by some stakeholders. Areas of consensus expressed across all participant groups were mostly focused on patient psychosocial support and access to medications.

**Conclusion:** This study was able to identify key perceptions elicited for an implementation strategy that combines peer medication delivery and health information technology to improve hypertension medication adherence. Pre-implementation stakeholder engagement can unearth unique perspectives around perceived benefits and challenges that can be used to refine strategies to increase the success of implementing evidence-based interventions in new contexts.

ClinicalTrials.gov identifier (NCT number): NCT05051124

## INTRODUCTION

Elevated blood pressure (BP) increases cardiovascular morbidity and mortality risk.^1–3^ Hypertension in the adult population has increased in recent years, with nearly 20 million people with hypertension residing in Sub-Saharan Africa.^4–6^ Nearly 60% of people living with hypertension in low-resource settings report suboptimal adherence to antihypertensive medications, contributing to uncontrolled hypertension.^5,7^ In Kenya, the prevalence of hypertension in the adult population is 23.7%, and hypertension treatment and control rates are suboptimal at 9% and 3%, respectively.^5^ Cost is among the primary factors that affect medication adherence, especially the indirect costs of transportation to access healthcare services.^8^

Some of the strategies identified that improve adherence and clinical outcomes in the management of hypertension in low-resource settings include improving medication possession and robust adherence monitoring systems.^9,10^ Peers have successfully been used to provide adherence counseling and psychosocial support for patients in the Academic Model Providing Access to Healthcare (AMPATH) healthcare delivery program.^8–11^ In addition, previous studies have shown that mobile group-based non-communicable disease (NCD) care, peer support, and improved medication supply chain strategies improve BP control.^11,12^ The use of health information technology (HIT), such as electronic medical record systems, mobile health (mHealth) approaches, and electronic prescribing, can also enhance medication adherence. ^13–15^ In low- and middle-income countries (LMICs), HIT has demonstrated beneficial impacts on clinical care processes for NCDs and can enhance peer support networks for NCD care.^17,18^

Each of the above components—peer support, medication delivery, and HIT—has been individually evaluated but not implemented in a combined and integrated fashion. Our study team therefore designed an implementation strategy that combines community-based medication delivery, peer support, and HIT, with the goal of improving medication adherence among patients with hypertension. The peers and technology for adherence, access, accountability, and analytics (PT4A) strategy aims to address the challenges of psychosocial barriers as well as medication possession that impact medication adherence. However, before implementation, we recognize that it is critical to engage key stakeholders (i.e., policymakers, clinicians, and community representatives) and anticipated beneficiaries (i.e., patients), to assess their perceptions of the planned implementation strategy, in order to ensure that implementation will be acceptable and feasible in our setting. The objective of this study, therefore, is to report on the perceptions of PT4A to optimize implementation.

## METHODS

### Study Setting

The study was conducted in western Kenya as part of the Academic Model Providing Access To Healthcare (AMPATH) program, an academic partnership between Moi University College of Health Sciences (MUCHS), Moi Teaching and Referral Hospital (MTRH), and a consortium of North American universities.^18^ AMPATH has established a robust chronic disease management (CDM) program that has provided clinical care to over 40,000 patients with hypertension across 69 health facilities in its catchment area in rural western Kenya.^11,18–20^ The multicomponent package includes task redistribution,^21^ clinical decision support using health information technology,^14,21^ consistent and secure medication supply,^14^ linkage and retention activities,^9^ community stakeholder engagement,^11,14,16^ and social support including microfinance groups.^16^

The CDM program employs the AMPATH medical records system (AMRS) infrastructure, which facilitates efficient and accurate capture and transmission of clinical information, decision support, and integration with a longitudinal electronic medical record.^21^ This infrastructure employs a customized version of OpenMRS, an open-source electronic medical record and informatics infrastructure that started in 2004 in Kenya and has now expanded to numerous countries worldwide.^11,18^ The clinical data allow information to be leveraged across clinical, public health, and research arenas simultaneously with the ultimate goal of improving care delivery.^5,18,21^

The three study sites were Webuye in Bungoma County, Kitale in Trans Nzoia County, and Turbo in Uasin Gishu County (**Figure 1**) in western Kenya. These three sites have a combined population of approximately 3.3 million people and a high burden of cardiovascular disease.^20,22^ The health facilities in these localities are in urban areas but are also utilized by the rural population in the surrounding areas.

**Figure 1.**
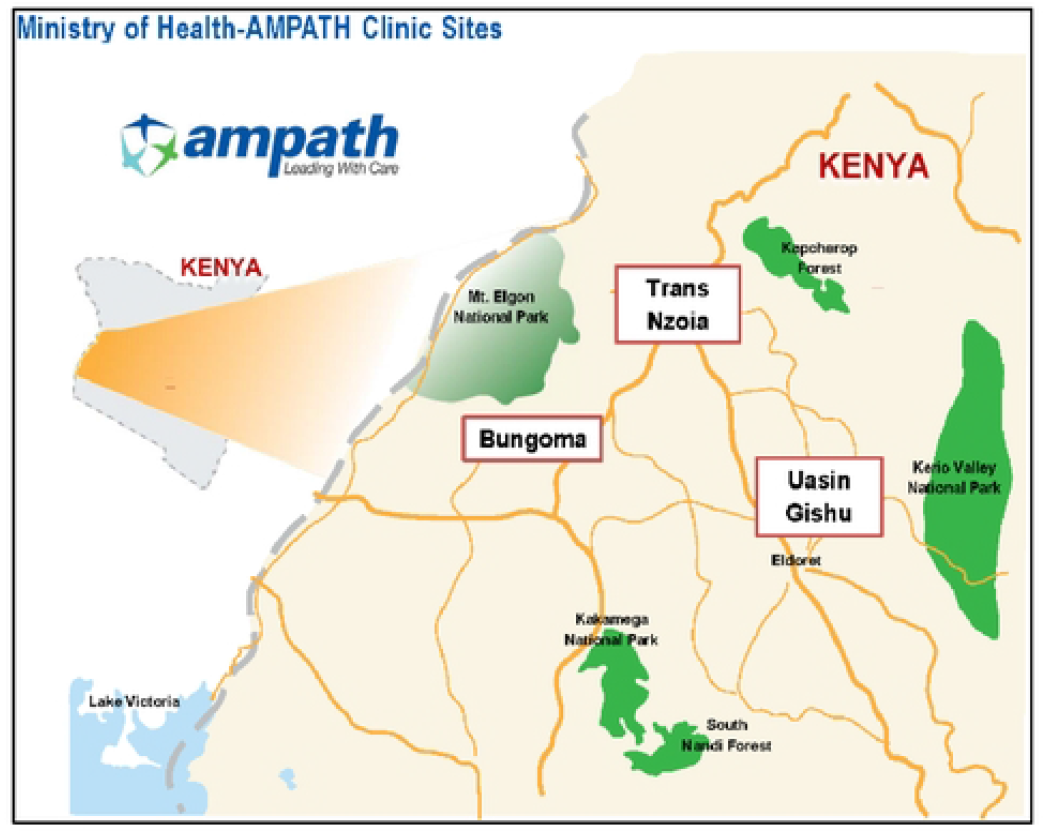
Map of Study Sites in the AMPATH Catchment Area of Western Kenya.

### Proposed PT4A implementation strategy

The planned PT4A implementation strategy integrates three components: peer coaching and support, community-based medication delivery, and HIT. These components of the implementation strategy are described below. **Figure 2** provides a visual display of how the PT4A implementation strategy aligns with the patient’s health facility visits and clinician encounters.

**Figure 2.**
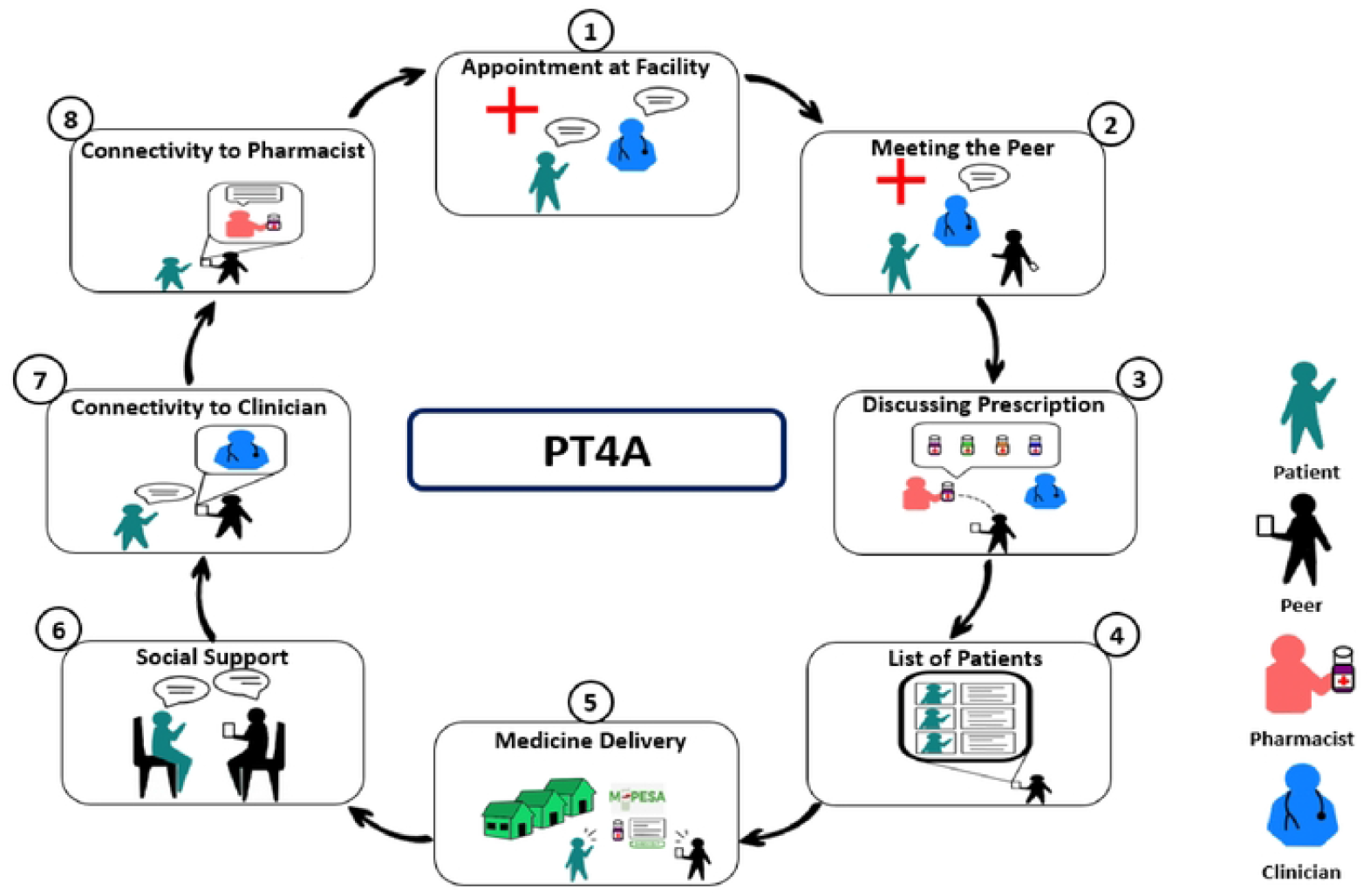
Visual Display of the PT4A Implementation Strategy. Steps in the process include: I. Patient is approached while at the clinic/facility, and they are informed about the implementation strategy. 2. Once the patient consents, they are introduced to the peer. 3. The peer and the patient meet,vith the pharmacist/pharmaceutical technologist to discuss the prescription and the refill plan. 4. The peer updates the list of participants requiring the medication deliveries. 5. The peer and the patient meet for medication delivery. 6. Medication adherence is assessed, and counseling support is given. 7 and 8. If additional consultation is required a teleconsultation is organized between the clinician and the patient and, if need be, the pharmacist. The patient is booked for a follow-up clinic visit, and the cycle continues.

#### Peer support

Peers would be lay individuals with shared disease experience who provide practical, social, emotional, and motivational assistance to patients. The peer-patient interaction would provide an opportunity for psychosocial support by the peer, with an emphasis on adherence assessment and counseling.

#### Medication Delivery

Medications would be delivered to patients with hypertension in the community who are enrolled in the PT4A program. Medication regimens would be packed by a qualified pharmacist or a pharmaceutical technologist, according to the patient’s prescription. Medications would be delivered for refills required in between the patient’s clinic visits, with the peer and patient agreeing on the exact delivery location in the community beforehand. If not enrolled in a health insurance plan that covers outpatient hypertension medications, patients would be expected to pay for medications through a mobile banking app prior to delivery. The goal of delivery would be to reduce transport costs to the health facility, avoid the opportunity cost of time lost from work, increase convenience for patients, and reduce facility congestion.

#### Health Information Technology (HIT)

The peer-patient interaction would be documented by the peer on a point-of-care HIT tool loaded onto an electronic tablet and built onto the existing AMRS infrastructure. The encounter form would include the delivery details, patient-reported medication adherence, pill count, and any challenges the patient reports to have been experiencing. The HIT tool would be pre-designed with decision support to aid in patient care, including providing reminders to provide adherence counseling if needed, and prompts to initiate a teleconsultation with the patient’s primary clinician if necessary. Finally, the tool would facilitate tracking and accountability by providing the peer with timely prompts when patient refills are nearly due, confirming receipt of medications by patients to improve accountability, and providing patient-level medication use analytics to predict future stock requirements for the pharmacy.

### Study Population and Sampling

We used qualitative research methods, including focus group discussions (FGDs) and key informant interviews (KIIs), to assess perceptions about the proposed PT4A implementation strategy. We conducted a total of 12 FGDs across the three study sites (counties), each with six to ten participants. In each county, a total of four FGDs were conducted: two FGDs with patients living with hypertension (separate for men and women), one FGD with clinic staff, and one FGD with community members. The local community members were recruited through the community mobilizers as guided by the AMPATH leadership; they included religious leaders, business leaders, village elders, and administrators. We also conducted four KIIs among health care administrators, one with the representative from the division of NCDs at the Kenya Ministry of Health and three with the county health representatives (one from each respective county). Health facility in-charges and county leadership assisted with contacting the participants who were eligible to participate in the KIIs.

Only participants above the age of 18 years who voluntarily gave written consent to participate in the study were included. Ethical approval was obtained from the Institutional Research and Ethics Committee (approval #0003812) at Moi University in Kenya and from the IRB at NYU Grossman School of Medicine (approval # i20-01579).

### Study Procedures

A set of question guides for the FGDs and KIIs was developed to explore the perceptions of the benefits and challenges of the different components of the PT4A implementation strategy. The question guide domains included the perceptions of peers contributing to health service provision, medication delivery, and psychosocial support (specific questions listed in Appendix A). The question guides were piloted in the MTRH hypertension outpatient clinic among both patients and the head of the division, which yielded useful feedback that was ultimately incorporated into the final question guides.

The FGD and KII sessions were conducted in English and Kiswahili by researchers with training and experience in conducting qualitative research. The KIIs and FGDs lasted approximately 60 and 90 minutes per session, respectively. The FGDs with community members and patients took place in conference centers, whereas the KIIs and the healthcare team FGDs were done at the facilities. All sessions were audio-recorded. For the FGDs, note takers also took notes on session proceedings. At the end of each session, participants were provided with reimbursement to cover their transport costs. All qualitative sessions were conducted between July and August 2021.

### Data analysis

The audio recordings of the sessions were transcribed verbatim and translated into English. The research team (CW, JW, BN, MK, and KK) verified the transcriptions and translations, then read through the transcripts and assigned preliminary codes. A codebook containing deductive codes was generated based on *a priori* themes such as perceptions of a peer-led project to support individuals living with hypertension, medication delivery at the community, and the use of a HIT tool to support peers in service delivery, as per the semi-structured interview guide. The research team then reviewed the final codes for validation and consensus. Each transcript was analyzed by one person (either CW, MK, or KK). Similar information was merged from the transcripts to create the major themes. A series of meetings was organized among the research team to reread, recode, and reanalyze, until consensus was achieved regarding the breadth and depth of emerging and conflicting themes and subthemes. NVIVO 12 qualitative data analysis software was used to organize, store, and analyze collected data.

## RESULTS

A total of 114 individuals were approached to participate in the study, out of whom 109 agreed to participate: 52 were patients with hypertension, 28 were community members, 25 were clinicians, and four were program/policy NCD planners. The primary reason for non-participation was due to personal obligations conflicting with the time required for interviews. Table 1 provides a breakdown of the demographic characteristics of study participants.

**Table 1:** Demographics of Study Participants.

| **Category** | **Patient FGDs** | **Clinician FGDs** | **Community Member FGDs** | **KIIs** |
| --- | --- | --- | --- | --- |
| **Participant Number** | 52 | 25 | 28 | 4 |
| **Mean Age in Years (±SD)** | 57.9 (±12) | 35.7 (± 6) | 45.8 (±10) | 47.3 (±11) |
| **Female Gender, n (%)** | 29 (56%) | 14 (56%) | 8 (29%) | 0 (0%) |
| **Maximum Formal Education Attained** |  |  |  |  |
| **None** | 6 | NA | 0 | 0 |
| **Primary** | 14 |  | 3 | 0 |
| **Secondary** | 20 |  | 12 | 0 |
| **Beyond Secondary** | 12 |  | 13 | 4 |
| **Average Monthly Income Category in Ksh** |  | NA |  | NA |
| **<1000** | 4 |  | 0 |  |
| **1000-2999** | 16 |  | 0 |  |
| **3000-4999** | 9 |  | 2 |  |
| **5000-9999** | 9 |  | 3 |  |
| **≥10000** | 14 |  | 23 |  |

### i. Fragility of trust: Safe spaces and shared experiences, but doubts about confidentiality

Community members and patients nearly universally viewed peers as approachable and friendly. Patients reported feeling safe and comfortable when opening up to peers about new symptoms and their shared experiences with hypertension treatment. From the perspective of the patients, meeting at more private locations closer to home facilitated honest and open discussion, which they believed would improve the monitoring and management of their condition.

> *“You become free. You can tell her anything. You can tell her how you feel”*. (Female FGD; patient living with hypertension).

In addition, patients also felt that peers truly cared for them and were committed to seeing them get well. Patients appreciated the compassion and care of someone consistently doing a follow-up on them while at home. Patients described peers as compassionate, caring, and having the best interest of patients at the forefront. This would create safe and non-judgmental spaces where challenges with hypertension management could be openly discussed. These perceptions increased the trust that patients felt toward peers, which would motivate them to continue with their treatment plans.

*“And then when I get medication, when she brings it to me, she explains to me well, at least she encourages me. The fact that she left the hospital to come here already gives me courage that I am supposed to continue adhering to medication*.*”* (Female FGD; patient living with hypertension)

The healthcare team and the administrators also perceived that shared experiences would facilitate social support, ultimately helping to overcome the challenges patients face with navigation of the health system and hypertension medication adherence. However, they expressed some doubts about peers’ ability to maintain the confidentiality of a patient’s private health information and responsibly handle said information. Some clinicians and administrators expressed concerns about peers not being well-trained in how to handle confidential information or possibly taking advantage of patients. While repeatedly mentioned by administrators and health workers, this concern was only expressed by a minority of patients and community members.

> “*There are those who do not keep quiet, privacy and you don’t know, they are not under any terms and conditions like maybe a health worker”* (FGD, HealthCare team).

### ii. Cautious optimism: Potential to improve hypertension service delivery, but persistent concerns

Across the board, participants felt that several components of PT4A had the potential to improve hypertension service delivery. Healthcare team members reported that peers would strengthen community-clinic linkage by helping to contact patients who missed clinic appointments.

*“But if there is a peer, defaulter tracing becomes easy. This person goes to the village and knows this person is in such a place and helps trace them*” (FGD, Healthcare team).

The HIT tool was felt to facilitate correct medication distribution and appropriate referrals, as well as aid in the improvement of health records management. In addition, patients and clinicians agreed that the HIT tool facilitated improved communication between peers, clinicians, and pharmacists, ultimately leading to better patient follow-up. Participants appreciated the teleconsultation component, which they felt would ensure that patients would receive the same clinical consultation as they would in person with minimal disruptions to their daily activities. The increased medication access and possession were also viewed favorably. All of these aspects were felt to facilitate the ability of the healthcare team to provide high-quality patient-centered care.

> *“For technology, it will enhance easier feedback between the patient and the clinician”* (FGD, Healthcare team).

These favorable impressions were counter-balanced by some concerns. Healthcare workers and administrators expressed concerns that communities might view peers as “*small doctors*” and prefer to consult them instead of trained medical personnel. In addition, both clinicians and community members expressed concern that there might be miscommunication of medical issues and potentially the spread of misinformation by peers to patients. Community members and patients questioned whether peers would have the requisite technology skills to operate the mobile device effectively or collect all the relevant data required during the peer-patient encounter, potentially adversely impacting the quality of care provided.

> *“Am just raising a concern that we should be cautious of overdependence of this patient to the peer*.” (FGD, HealthCare team).

Some community members cited poor road infrastructure, which could limit transportation and potentially decrease the efficiency of peer delivery in some areas. In addition, community members and patients expressed concerns about logistical issues, such as the patients needing to have phones for communication with their peers and ensuring sufficient network coverage, both of which could be difficult and expensive to maintain for many individuals in the area.

> *“These phones also have issues with network. There are some areas that might not have network. So, communication there can be a barrier, and also money. If it is the peer’s phone, if the airtime runs out, the conversation will end there. If it is a video call, it will end there so the patient wouldn’t have been helped”* (FGD, Community Member).

### iii. Owning our healthcare: Increased community awareness and engagement in hypertension management

All participants believed that the PT4A implementation strategy would create more awareness about hypertension management and address misconceptions about the disease, thus reducing the associated stigma of chronic diseases. They also anticipated that PT4A would enhance community participation in managing their health issues.

Administrators, in particular, felt that engaging peers from the community would create a sense of confidence that hypertension could be managed at the community level while promoting the active engagement of patients in their care. In addition, community members would be able to hold peers accountable for their assigned duties. As a result, this would promote community ownership of hypertension management.

“*Then there is that ownership, we are owning our own health problems. You see using peer educators is empowering the community to have their own resource person, whom they know, whom they trust, whom they respect and whom they have confidence in rather than health workers going there who doesn’t know the culture, the norms of that society and do’s and don’ts”* (KII, administrator).

Community members felt that having a peer in the community would encourage individuals living in the community to participate in screening and health promotion activities since a greater number of individuals would become more aware of the condition. Patients also felt that PT4A would create a more supportive and understanding environment since other community members would now have more understanding of and empathy for their condition and associated challenges. This supportive environment would encourage those who might otherwise fear stigma, discrimination, or the consequences of the condition.

*“When I know my peer is coming, I will inform my neighbors that I have a visitor. When she comes, she might even advise those who don’t have pressure and they will understand*.*”* (FGD, female patient living with hypertension)

## DISCUSSION

In this qualitative study from western Kenya, our proposed implementation strategy of integrated peer-based medication delivery and health information technology was perceived to potentially improve community-based hypertension management, medication possession, and patient-level adherence, leading to improved health outcomes. Stakeholders thought that the implementation strategy could create safe spaces for people living with hypertension to openly discuss shared disease experiences, enhance trust, lead to more effective and efficient service delivery, and promote community ownership of hypertension care. However, they were also concerned about issues related to peers maintaining patient confidentiality, peers operating outside their scope of work, peer capability to deliver on their expected roles, and environmental logistical challenges hindering peer medication delivery.

While there was consensus agreement on several themes across the different participant groups (patients, healthcare workers, community members, and administrators), there were some notable differences in perceptions as described in detail above and illustrated in **Figure 3**. This diversity of perspective reinforced the importance of recruiting a wide variety of participants across several different participant categories. Triangulation of perceptions expressed by a diverse group participants ultimately will lead to a final implementation strategy that is more acceptable to a larger group of stakeholders, thus enhancing likelihood of implementation success.

**Figure 3:**
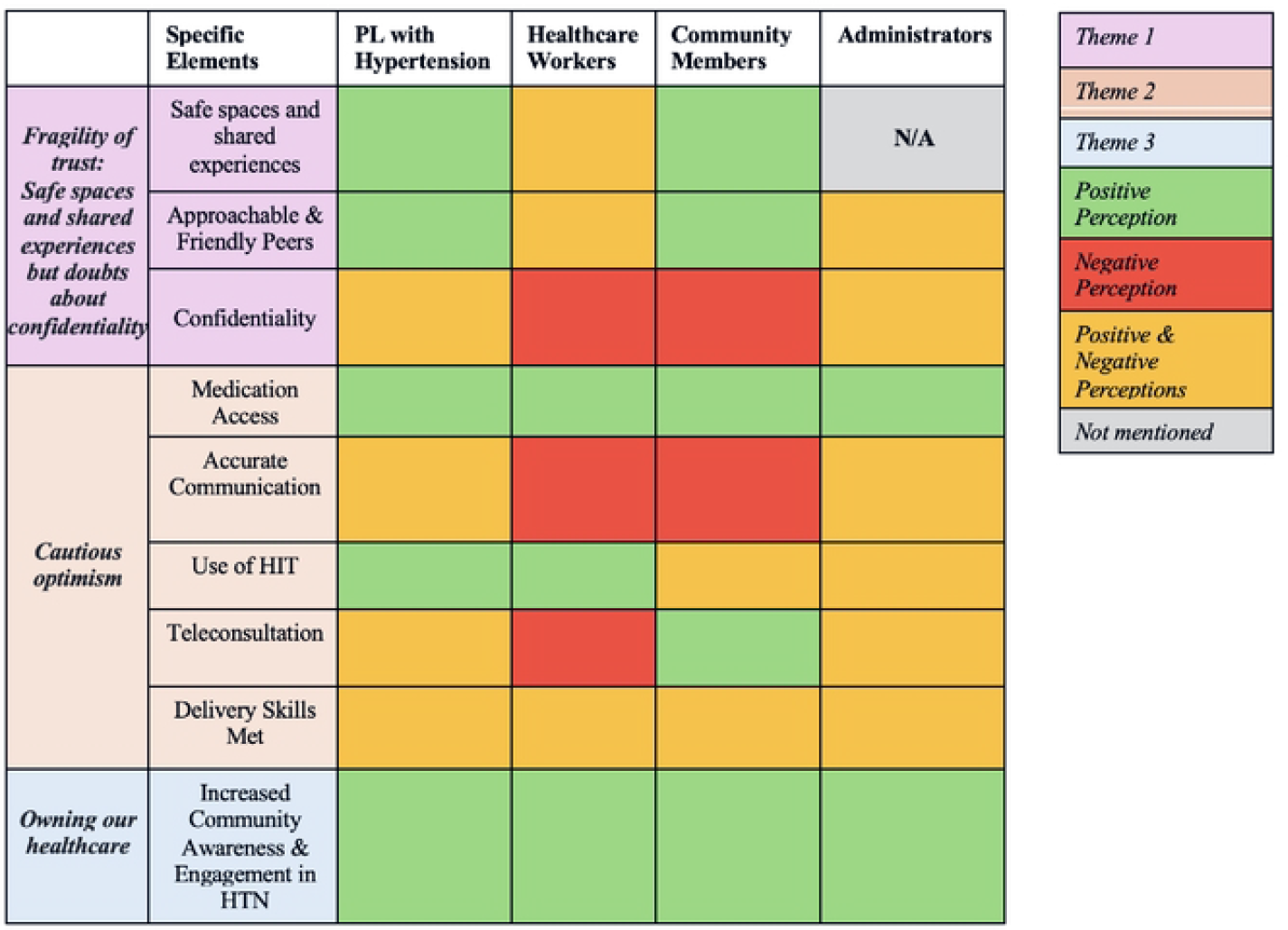
Thematic Variation Across Patients, Healthcare Workers, Community Members, and Administrators. This figure presents a color-coded table summarizing ho,v different stakeholder groups perceive specific elements related to hypertension (HTN) care and engagement. The table is organized by three overarching themes: (l) Fragility of trust [purple] (2) Cautious optimism [tan], and (3) Owning our healthcare [blue]. The columns represent the four different groups whose perceptions,vere assessed: (1) People Living,vith Hypertension [PL v,,ith HTNJ, (2) Healthcare Workers, (3) Community Members, and (4) Administrators. Color coding indicates the nature of their perceptions: green indicates positive perceptions, red arc negative, yellow represents mixed perceptions, and grey indicates unavailable information

Peers were generally perceived as trustworthy because they were viewed as approachable and friendly, had shared lived experiences with hypertension, and had the patient’s best interests at heart. Trust is fundamental to effective interpersonal relations and community living, and is an important aspect of people’s experiences with both the health system and patient-clinician interactions.^23–26^ Consistent with other studies, trust is typically associated with high-quality communication and interaction, which facilitates disclosure by the patient, enables the practitioner to encourage necessary behavior changes, and permits the patient to have greater autonomy in decision-making about treatment.^24,25^

Trust is traditionally viewed as consisting of three key components: benevolence, capability, and integrity.^27^ Benevolence was reflected in patients’ perceptions that peers would align with their interests and would work with compassion and care. Peers have a lived experience with the health condition and have also experienced social challenges, which allows them to listen without judgment and the potential to give hope to those living with hypertension. They felt that this would create a safe space to openly discuss any challenges they were facing with disease management. Strong benevolence-trust relationships have been shown to improve adherence to treatment plans, and prior work has identified benevolence as a key factor of interpersonal trust especially between patients and providers.^23, 24^ For instance, a peer-support program for hypertension treatment in Iran demonstrated the importance of this aspect of trust-benevolence and how peers’ relationships with patients can greatly improve adherence to not only medication regiments but also diet and physical activity changes.^27, 29^ Our results align with these previously reported findings.

With respect to the capability component of trust, healthcare workers expressed some negative perceptions regarding peers in this regard. In particular, they were worried that peers may not be equipped to handle the HIT tool and complete the technology-dependent tasks. These valid concerns highlight the crucial role of establishing and maintaining continuous peer training and education programs geared to emphasizing correct HIT use and computer skills for effective patient encounter documentation and basic aspects of hypertension management in which peers would be expected to be involved. Previous work has shown that proper training and resources can help to address concerns regarding limited medical and technical knowledge of peers.^26^

Healthcare workers were also concerned about the peers’ integrity with respect to maintaining patient confidentiality and the protection of sensitive personal information. Other studies have supported the idea that patients expect members of their health team to respect confidentiality, this being an important aspect of integrity.^30,31^ Administrators felt that recruiting peers from their own respective communities, where they would be expected to work, would allow existing community ties to be leveraged to hold peers accountable to their expected duties, which could help to maintain peer integrity. In addition, establishing clear roles and expectations, as well as professional and ethical standards regarding patient confidentiality and private health information, would further enhance integrity and aid in alleviating some of those concerns.^29^

An important aspect was that both peers and the HIT component were viewed as helping to increase community-clinic linkage and improve overall care delivery. Peers can advocate for the needs of patients and communities, acting as mediators between them and the healthcare system to render care more readily accessible and adaptable to patient needs.^30^ Patients would now have increased follow up even while geographically remaining in the community/village. The data collected during patient home visits would help the healthcare personnel be able to review if patients are managing the condition sufficiently, as well as any particular issues or needs expressed by the patients. Other studies are also increasingly demonstrating that the use of HIT can improve the quality and coordination of care and lead to better health outcomes.^31,32^ Digital health technology allows for data collection, decision support, tele-consultation, and adherence counseling while in the community setting, and is therefore akin to an extension of the health system while staying in the community.^33^ In addition, strengthening community-clinic linkage can help to increase trust in the health system and reduce associated obstacles in accessing care.^27,28^

The potential for increased community engagement and activation was also perceived as beneficial to patients. Incorporating peers who reside within the existing community can work to potentially reduce stigma surrounding hypertension and its managements by normalizing conversations about the disease and treatments within social networks that are familiar to patients. Prior studies of similar peer support networks have shown that when counseling occurs between those who already have trusting relationships, this can help individuals feel less isolated and encourage them to be more open about their condition.^27^ Peers acting as community-based resources can also strengthen accountability in the local community along with ownership of hypertension management. This is consistent with previous evidence demonstrating that communities actively participating in disease management can increase interaction with healthcare professionals, support sustained behavioral and lifestyle changes, and improve individual health outcomes.^31,34^

We acknowledge the following limitations. Our qualitative approach relies on participants’ responses that might be specific to their life experiences, prior knowledge, or biases. Thus, the responses and subsequent thematic analysis may not reflect the entirety of the population or other communities outside of western Kenya. We attempted to mitigate this limitation by enrolling a diversity of participants representing a variety of stakeholder groups. However, we acknowledge that we could have recruited an even wider participant group such as other healthcare workers, local village elders/chiefs, or even family and friends of patients. Second, the geographic setting was admittedly quite small and restricted to western Kenya, adding to the challenge of generalizability. Finally, the research project was undertaken in a region where the health care infrastructure has been extensively developed over time, and we recognize that this might be notably different from other settings where such health system investments have not occurred. Despite this, we feel that the themes that arose from this qualitative exploration have relevance for individuals and communities living in low-resource settings worldwide.

## CONCLUSION

The findings of this study provide critical information that will be used to refine and finalize the planned PT4A implementation strategy. We were able to uncover opportunities to address important implementation issues to maximize the success of the ultimate implementation strategy. These results emphasize the importance of conducting similar pre-implementation research in other settings. Pre-implementation stakeholder engagement representing a diverse array of perspectives can unearth unique insights about perceived benefits and challenges that can be used to refine strategies and increase the success of implementing evidence-based interventions in new contexts.

## Data Availability

Data scrubbed of all identifiers will be made available upon request.

https://osf.io/hvpqa/overview?view_only=8ebeaa8b1fce4978a53c45f30244ecba

